# Exposure to valsartan products containing nitrosamine impurities in the US, Canada, and Denmark

**DOI:** 10.1101/2023.12.05.23299542

**Authors:** Efe Eworuke, Mayura U. Shinde, Laura Hou, J. Michael Paterson, Peter Bjødstrup Jensen, Judith C. Maro, Ashish Rai, Anton Pottegård, Daniel Scarnecchia, Yuanling Liang, Deborah Johnson, Robert W. Platt, Hana Lee, Marie C. Bradley

## Abstract

**Background:** Following the mass recall of valsartan products with nitrosamine impurities in July 2018, the number of patients exposed to these valsartan products, the duration of exposure, and the potential for cancer remains unknown. Therefore, we assessed the extent and duration of use of valsartan products with a nitrosamine impurity in the US, Canada, and Denmark.

**Methods:** We conducted a retrospective cohort study using administrative healthcare data from the US FDA Sentinel System, four Canadian provinces that contribute to the Canadian Network for Observational Drug Effect Studies (CNODES), and the Danish National Prescription Registry. Patients, 18 years and older between May 2012 and December 2020 with a valsartan dispensing were identified in each database. Patients were followed from the date of valsartan dispensing until discontinuation. We defined four valsartan exposure categories based on nitrosamine impurity status; recalled generic products with confirmed NDMA/NDEA levels (recalled-tested); recalled generic products that were not tested (recalled); non-recalled generic and non-recalled branded products. In Denmark, recalled-tested category was not included due to absence of testing data. The proportion and duration of use of valsartan episodes stratified by nitrosamine-impurity status.

**Results:** We identified 3.3 and 2.8 million (US) and 51.3 and 229 thousand (Canada) recalled-tested and recalled valsartan exposures. In Denmark, where valsartan exposure was generally low, there were 10,747 recalled exposures. Immediately after the recall notices were issued, there was increased rates of switching to a non-valsartan ARB. The mean duration of use of the recalled-tested products was 167(±223.1) and 146(±255.8) days in the US and Canada respectively. For the recalled products, mean cumulative duration of use was 178(±249.6), 269(±397.3) and 166(±251.0) days in the US, Canada, and Denmark, respectively.

**Conclusion:** In this cohort study, despite widespread use of recalled generic valsartan between 2012 and 2018, the duration of use was relatively short and likely did not pose an elevated risk of nitrosamine-induced cancer. However, since products with nitrosamine impurity could have been on the market over a six-year period, patients potentially exposed to these products for longer duration could have a different risk of cancer.

## INTRODUCTION

The mass recall of valsartan products with nitrosamine impurities in July 2018 prompted a series of investigations into the etiology and level of nitrosamine impurities, and the potential for cancer risk. Regulatory agencies quickly learned that the presence of N-nitrosodimethylamine (NDMA) and N-nitroso-diethylamine (NDEA) impurities in some valsartan products occurred as far back as 2012^1–5^ due to a change in manufacturing processes. Our previous study^6^ investigated trends in valsartan and other ARB use and found that the first recall notice for valsartan resulted in substantial decline in use due to increased switching to other ARBs. However, the extent of exposure to valsartan products with nitrosamine impurities before the recall, and the associated potential cancer risk remains unknown. In October 2018, regulatory agencies published test results showing NDMA levels in recalled valsartan products exceeded safe levels.^6,7^ The results were subsequently updated in February 2019 with the NDEA levels. For reference, consuming up to 0.096 micrograms (mcg) of NDMA or 0.0265 mcg of NDEA per day is considered reasonably safe for human ingestion based on lifetime exposure. The highest NDMA levels were between 14 and 20 mcg, while the highest NDEA levels were between 1.1 and 1.3mcg. Since cancer risk is dependent on both dose and years of exposure, it was determined that if 8,000 patients took the maximum recommended daily dose (320mg daily) of valsartan containing NDMA at levels of 96 ng/day (0.096 mcg) and 18,000 people took the highest dose of valsartan containing NDEA at levels of 26.5 ng/day (or 0.0265 mcg/day) for four years, there may be only one additional cancer case.^7^ This study examined the extent and duration of use of valsartan products with nitrosamine impurities and estimated the potential for cancer based on the observed duration of use of these products in the US, Canada, and Denmark.

## METHODS

A retrospective cohort study was conducted between January 1, 2012, through December 31, 2020, using data from six data partners that contribute data to the US FDA Sentinel System, data from four Canadian provinces that contribute to the Canadian Network for Observational Drug Effect Studies (CNODES), and the Danish National Prescription Registry. The FDA’s Sentinel system includes data on medical encounters (inpatient and outpatient diagnoses and procedures) and outpatient pharmacy claims data (retail and mail order filled prescriptions) accrued during health plan enrollment periods. These data are routinely quality-assured and transformed into the Sentinel Common Data Model to facilitate queries.^12^ Additional descriptions of data sources are provided in Supplement A.

We included patients aged 18 years and older with at least one valsartan dispensing during the study period, between January 2012 to December 2020 for US and Canada and January 2012 to May 2020 for Denmark. Four valsartan exposure categories were defined based on valsartan impurity status; recalled products with a laboratory test result confirming the presence of NDMA/NDEA based on FDA and Health Canada analyses (recalled-tested); recalled products that were not tested (recalled); non-recalled generic and non-recalled branded valsartan-containing products (Diovan, Entresto, Exforge and Sandoz-Valsartan). In Denmark, we did not have access to NDMA/NDEA testing data, hence valsartan products were categorized into recalled, non-recalled generic and non-recalled branded categories. The algorithms for exposure definitions are presented in supplement B. Valsartan exposure episodes were created by summing the number of days dispensed with a gap of 30 days or less for the respective exposure category. Patients were followed until they discontinued the product, switched to another valsartan exposure category or a different angiotensin receptor blocker (ARB), disenrollment, death, or study end date. We calculated the proportion of valsartan episodes as the number of valsartan episodes in each exposure category divided by the total valsartan episodes each quarter per year. We also calculated the rate of switching, defined as the number of valsartan episodes for each exposure category that ended in a switch, divided by the total valsartan episodes each quarter per year. Lastly, we estimated the length of episode duration by calculating mean, median and quartiles (lowest and highest median, 25^th^ and 75^th^ percentile recorded across the data partners) between May 2012 and December 2018 (period when we assumed valsartan products with NDMA/NDEA impurity were on the market) for each recall category. All source data were transformed into the Sentinel common data model^11^, and analyzed using Sentinel’s Query Request Program v10.1.1 with custom coding, allowing for the deployment of a common analytical program in all countries.

This study conducted in the FDA’s Sentinel System is a public health surveillance activity conducted under the authority of the FDA and, accordingly, is not subject to Institutional Review Board oversight.^9–11^ This study followed the Strengthening the Reporting of Observational Studies in Epidemiology (<u>STROBE</u>) reporting guideline.

## RESULTS

During the study period (May 2012-Decemeber 2020), we identified 7,925,941; 442,189 and 16,260 valsartan episodes in US, Canada, and Denmark. The distribution of valsartan use by exposure category during the study period for all three countries is presented in Table 1. Exposure to both recalled-tested and recalled products was extensive in the US and Canada. We identified 3.3 (41.6%) and 2.8 (35.7%) million recalled-tested and recalled valsartan exposure episodes in the US and 51,315 (11.6%) and 291,229 (65.9%) recalled-tested and recalled episodes were identified in the Canadian database. In Denmark, the proportion of recalled valsartan exposure was high; 10,747 (66.1%) recalled episodes were identified. Exposure to non-recalled generic and non-recalled branded valsartan products was comparable to the exposure to the recalled products, for all three countries. Around 3.3 (41.5%) and 3.2 (40%) million non-recalled generic and branded episodes, respectively, were identified in Sentinel, while 35,978 (8.1%) and 215,999 (48.8%) episodes for non-recalled generic and branded were identified in the Canadian data. In Denmark, 9,395 (57.8%) and 7,632 (46.9%) non-recalled generic and branded valsartan episodes were identified (Table 1).

**Table 1.**
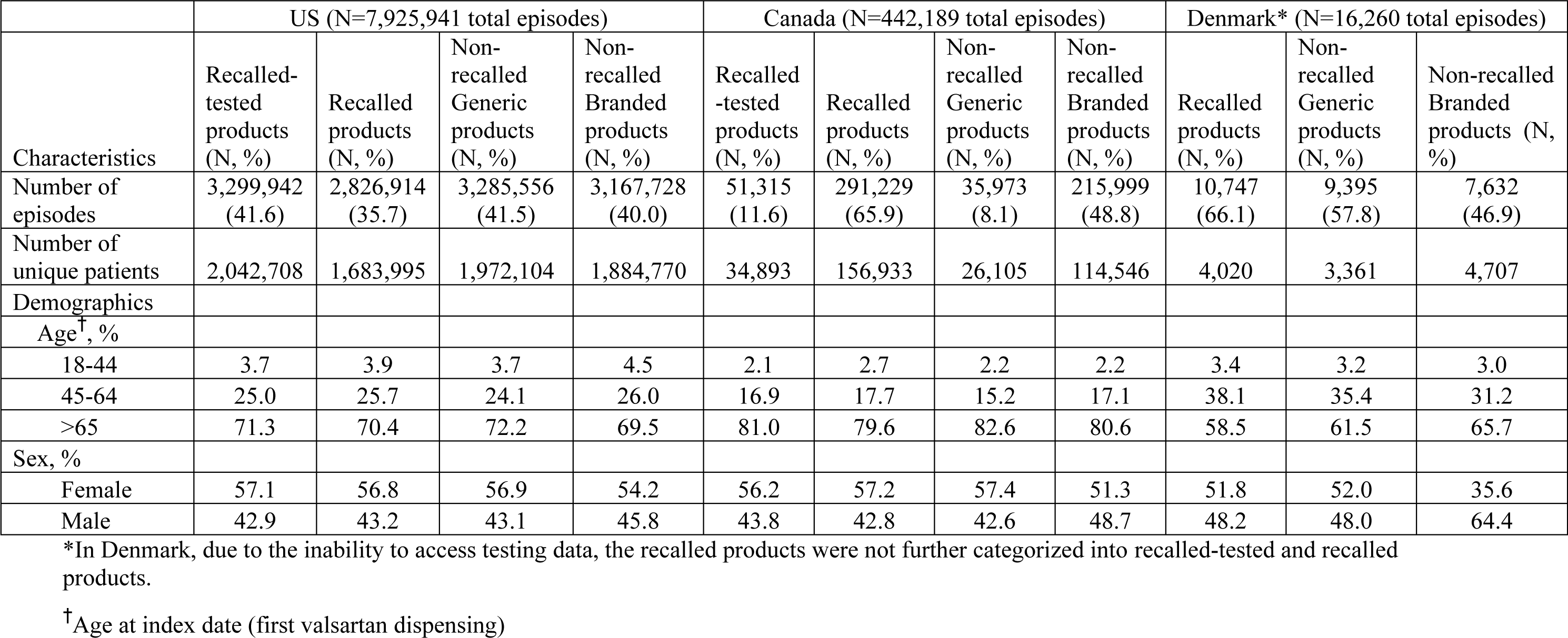
Characteristics of valsartan users, stratified by valsartan nitrosamine-impurity status for US, Canada, and Denmark between May 1, 2012, through December 31, 2020.

|  | US (N=7,925,941 total episodes) |  |  |  | Canada (N=442,189 total episodes) |  |  |  | Denmark* (N=16,260 total episodes) |  |  |
| --- | --- | --- | --- | --- | --- | --- | --- | --- | --- | --- | --- |
| Characteristics | Recalled-tested products (N, %) | Recalled products (N, %) | Non-recalled Generic products (N, %) | Non-recalled Branded products (N, %) | Recalled -tested products (N, %) | Recalled products (N, %) | Non-recalled Generic products (N, %) | Non-recalled Branded products (N, %) | Recalled products (N, %) | Non-recalled Generic products (N, %) | Non-recalled Branded products (N, %) |
| Number of episodes | 3,299,942 (41.6) | 2,826,914 (35.7) | 3,285,556 (41.5) | 3,167,728 (40.0) | 51,315 (11.6) | 291,229 (65.9) | 35,973 (8.1) | 215,999 (48.8) | 10,747 (66.1) | 9,395 (57.8) | 7,632 (46.9) |
| Number of unique patients | 2,042,708 | 1,683,995 | 1,972,104 | 1,884,770 | 34,893 | 156,933 | 26,105 | 114,546 | 4,020 | 3,361 | 4,707 |
| Demographics |  |  |  |  |  |  |  |  |  |  |  |
| Age<sup>†</sup>, % |  |  |  |  |  |  |  |  |  |  |  |
| 18-44 | 3.7 | 3.9 | 3.7 | 4.5 | 2.1 | 2.7 | 2.2 | 2.2 | 3.4 | 3.2 | 3.0 |
| 45-64 | 25.0 | 25.7 | 24.1 | 26.0 | 16.9 | 17.7 | 15.2 | 17.1 | 38.1 | 35.4 | 31.2 |
| >65 | 71.3 | 70.4 | 72.2 | 69.5 | 81.0 | 79.6 | 82.6 | 80.6 | 58.5 | 61.5 | 65.7 |
| Sex, % |  |  |  |  |  |  |  |  |  |  |  |
| Female | 57.1 | 56.8 | 56.9 | 54.2 | 56.2 | 57.2 | 57.4 | 51.3 | 51.8 | 52.0 | 35.6 |
| Male | 42.9 | 43.2 | 43.1 | 45.8 | 43.8 | 42.8 | 42.6 | 48.7 | 48.2 | 48.0 | 64.4 |

\*In Denmark, due to the inability to access testing data, the recalled products were not further categorized into recalled-tested and recalled products.
<sup>†</sup>Age at index date (first valsartan dispensing)

### Trends in valsartan use

The trends in valsartan use, stratified by valsartan impurity status for US, Canada and Denmark are presented in Figures 1-3.

**Figure 1.**
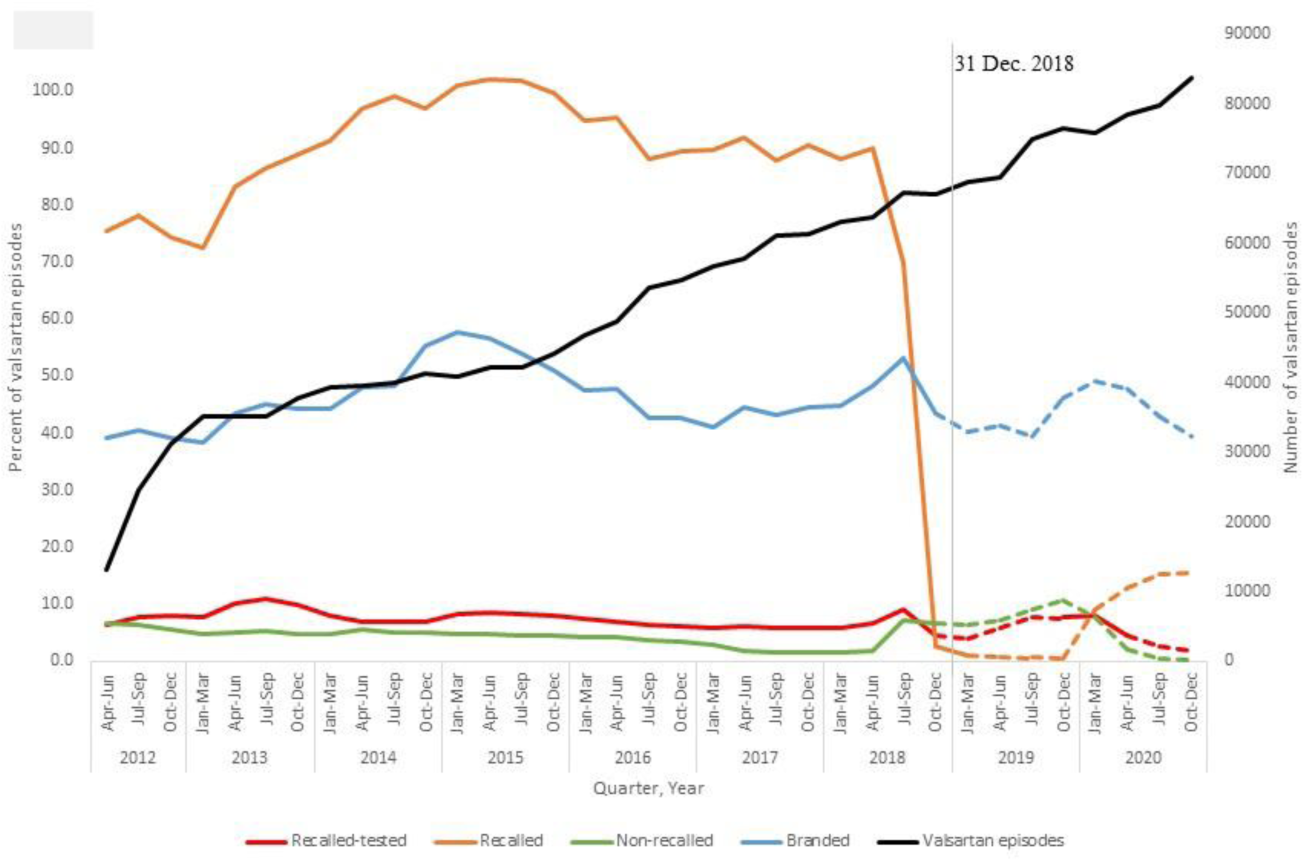
Trends in valsartan utilization stratified by nitrosamine impurity status in the US. The solid lines represent valsartan use while the products with nitrosamine impurities were on the market; the dotted lines represent use when the products with nitrosamine impurities were assumed to be removed from the market (Jan 2019-Dec 2020). For recalled products, use after 2019 reflects production without nitrosamine impurities.

**Figure 2.**
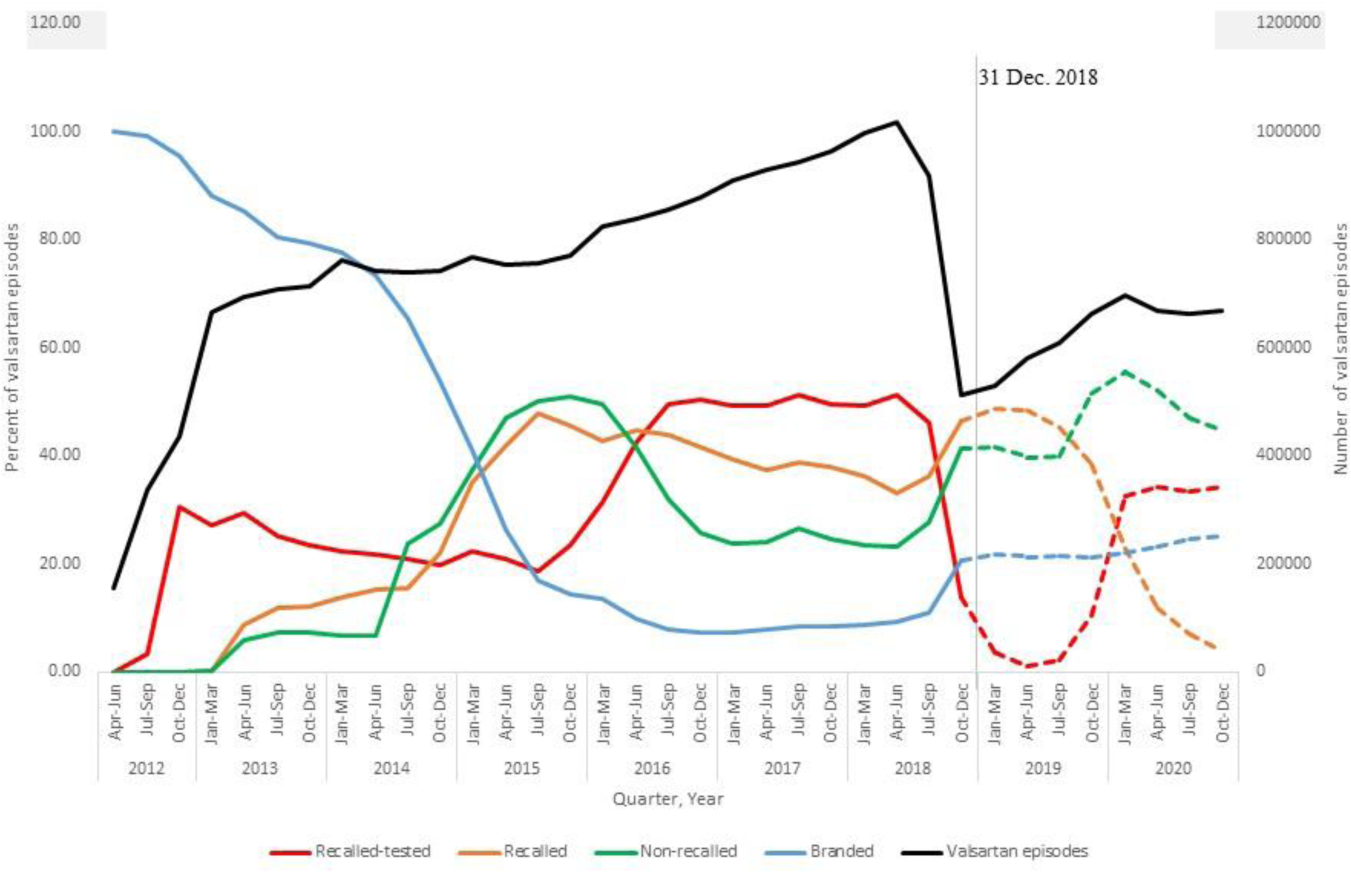
Trends in valsartan utilization stratified by nitrosamine impurity status in Canada. The solid lines represent valsartan use while the products with nitrosamine impurities were on the market; the dotted lines represent use when the products with nitrosamine impurities were assumed to be removed from the market (Jan 2019-Dec 2020). For recalled products, use after 2019 reflects production without nitrosamine impurities.

**Figure 3.**
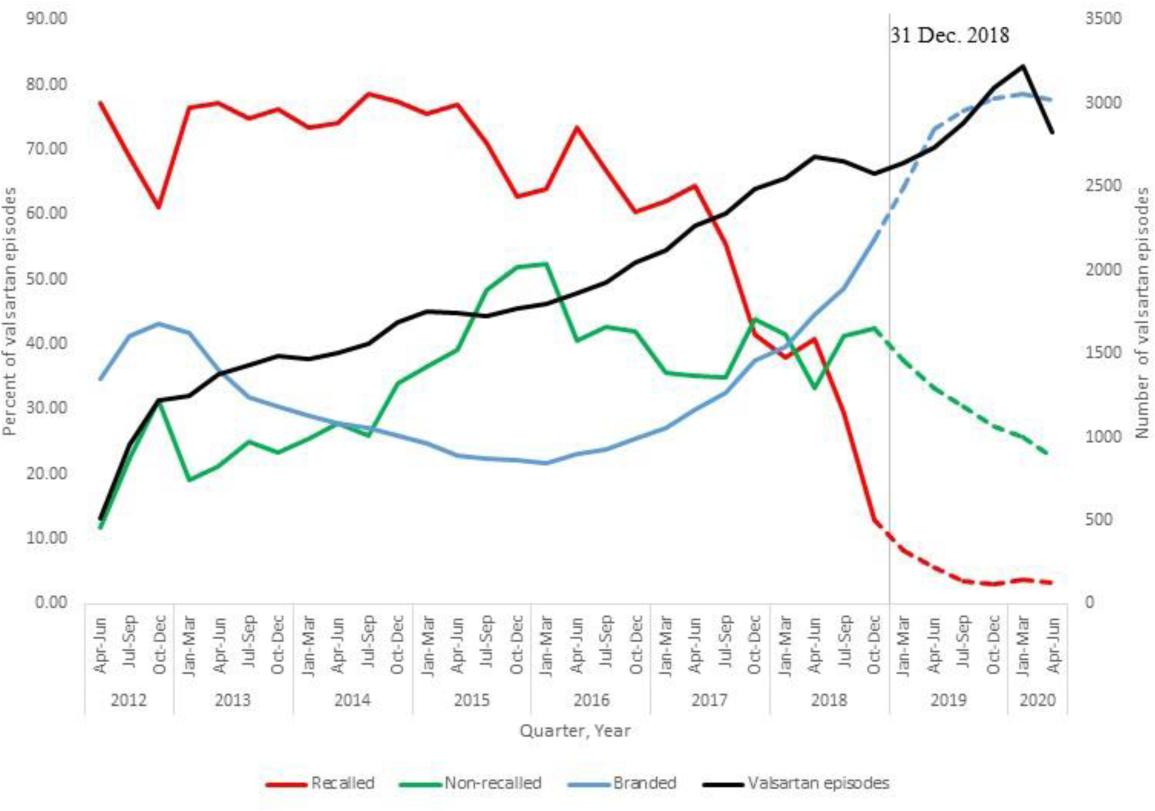
Trends in valsartan utilization stratified by nitrosamine impurity status in Denmark. The solid lines represent valsartan use while the products with nitrosamine impurities were on the market; the dotted lines represent use when the products with nitrosamine impurities were assumed to be removed from the market (Jan 2019-Dec 2020).

#### US

In the US, between 2012 and 2015, we observed a steady decline in use of valsartan branded episodes, along with a corresponding increase in the share of the generic (recalled-tested, recalled and non-recalled) products (Figure 1). Recalled-tested products had the largest share of the generic valsartan market in 2012; however, between 2014 and 2015, both non-recalled and recalled products equally shared the valsartan market (Figure 1). Between the third calendar quarter (Q3: July-September) of 2016 and Q3-2018, recalled-tested valsartan products had the largest market share of valsartan products, followed by the recalled products. From Q3 to Q4 (October-December) 2018, we observed a steep drop (from 46.0% to 13.9%) in the proportion of recalled-tested valsartan products. The decline in use of recalled products started from Q1 (January-March) 2019.

#### Canada

In Canada, recalled products were the most dispensed valsartan products, followed by non-recalled branded products (Figure 2). Between Q3- and Q4-2018, there was a steep drop in the proportion of recalled products (from 90% in Q2 (April-May) 2018 to 2.6% in Q4-2018); while the trends for non-recalled branded products remained unaffected. Use of recalled-tested and non-recalled generic products was low during the study period.

#### Denmark

In Denmark, recalled products were the most dispensed valsartan products (Figure 3). Decline in the use of these products beginning in 2017 coincided with the steady increase in non-recalled branded products. When the recall notice was issued (Q3-2018), there was a steep decline from 41% (Q3-2018) to 13% (Q4-2018) in recalled products, whereas the proportion of non-recalled branded products increased from 49% to 56% around the same time.

### Duration of valsartan episodes

The duration of valsartan exposure episodes, stratified by valsartan-impurity status for the three countries, is presented in Table 2. The mean duration of use of the recalled-tested products was 166.9(±223.1) and 145.5(±255.8) days in the US and Canada. The 75^th^ percentile ranged from 60-222 days for US and 139-192 days for Canada. For the recalled products, mean duration of use was 178.3(±249.6), 269.0(±397.3) and 166(±251.0) days for US, Canada, and Denmark. The 75^th^ percentile range was 60-230, 315-407 and 181 days for US, Canada, and Denmark. The mean duration of use for the non-recalled generic products appeared comparable to mean duration for recalled-tested (US, Canada) and recalled products (Denmark). In Canada (319.2 (±489.1)) and Denmark (321.6(±376.2) days), the mean duration of use of the non-recalled branded products was much longer compared to the duration of use in the US (167.7(±228.0) days).

**Table 2.**
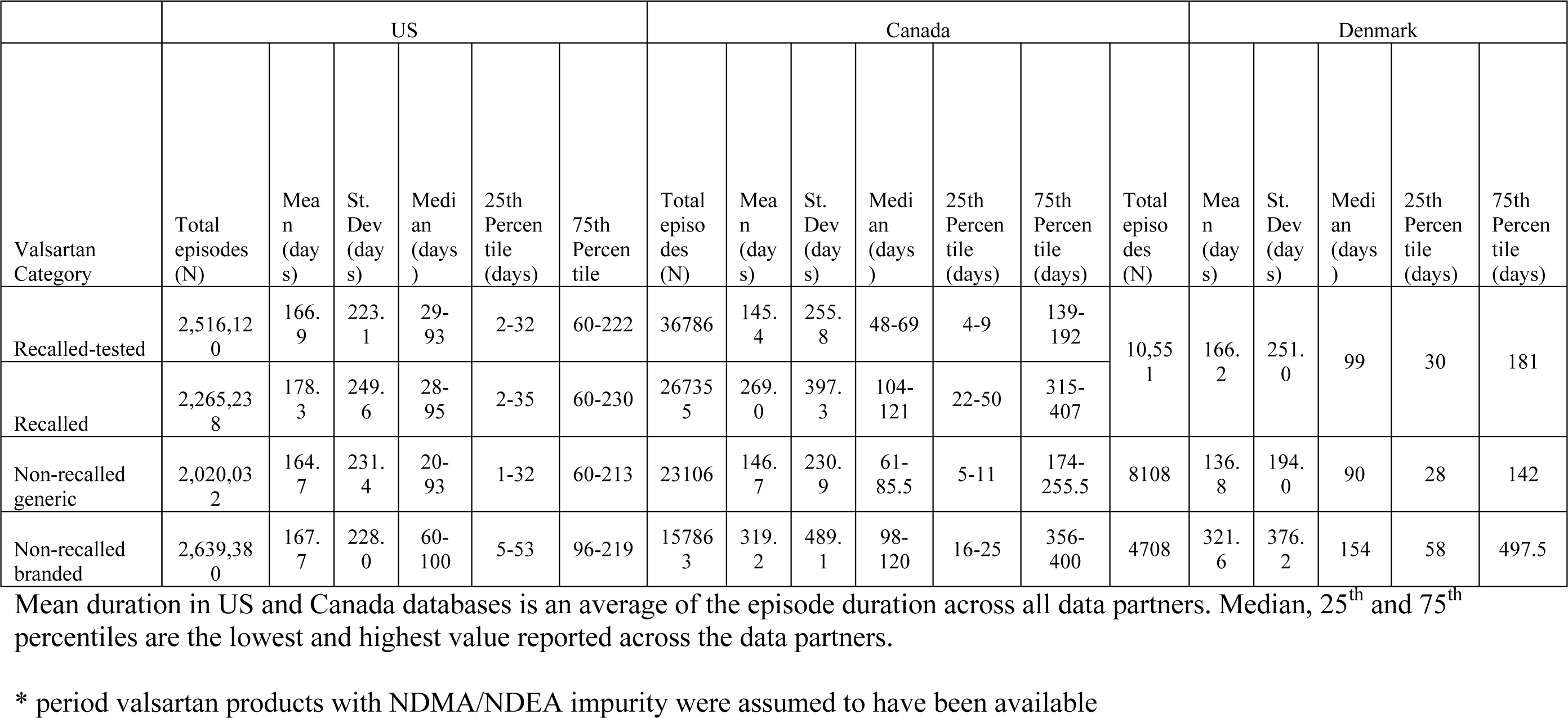
Summary statistics for the duration of valsartan episodes between May 1, 2012, and December 31, 2018*, by valsartan nitrosamine-impurity status.

|  | US |  |  |  |  |  | Canada |  |  |  |  |  | Denmark |  |  |  |  |  |
| --- | --- | --- | --- | --- | --- | --- | --- | --- | --- | --- | --- | --- | --- | --- | --- | --- | --- | --- |
| Valsartan Category | Total episodes (N) | Mean (days) | St. Dev (days) | Median (days) | 25th Percentile (days) | 75th Percentile | Total episodes (N) | Mean (days) | St. Dev (days) | Median (days) | 25th Percentile (days) | 75th Percentile (days) | Total episodes (N) | Mean (days) | St. Dev (days) | Median (days) | 25th Percentile (days) | 75th Percentile (days) |
| Recalled-tested | 2,516,120 | 166.9 | 223.1 | 29-93 | 2-32 | 60-222 | 36786 | 145.4 | 255.8 | 48-69 | 4-9 | 139-192 | 10,551 | 166.2 | 251.0 | 99 | 30 | 181 |
| Recalled | 2,265,238 | 178.3 | 249.6 | 28-95 | 2-35 | 60-230 | 267355 | 269.0 | 397.3 | 104-121 | 22-50 | 315-407 |  |  |  |  |  |  |
| Non-recalled generic | 2,020,032 | 164.7 | 231.4 | 20-93 | 1-32 | 60-213 | 23106 | 146.7 | 230.9 | 61-85.5 | 5-11 | 174-255.5 | 8108 | 136.8 | 194.0 | 90 | 28 | 142 |
| Non-recalled branded | 2,639,380 | 167.7 | 228.0 | 60-100 | 5-53 | 96-219 | 157863 | 319.2 | 489.1 | 98-120 | 16-25 | 356-400 | 4708 | 321.6 | 376.2 | 154 | 58 | 497.5 |

Mean duration in US and Canada databases is an average of the episode duration across all data partners. Median, 25<sup>th</sup> and 75<sup>th</sup> percentiles are the lowest and highest value reported across the data partners.
\* period valsartan products with NDMA/NDEA impurity were assumed to have been available

### Valsartan switching patterns

In US and Canada, we observed relatively stable switching patterns for both recalled-tested and recalled products prior to Q2-20718 (Supplemental Figures 1 and 3). In Q2-2018, there was an immediate increase in the switching between valsartan exposure categories and to a non-valsartan ARB. In Denmark, a similar increase in switching was also observed in Q2-2018; however, there was frequent switching from recalled products to non-recalled valsartan products (Supplemental Figure 5). Interestingly, in Q2-2018, there was increased switching from non-recalled valsartan products to a non-valsartan ARB in the US and Canada (Supplemental Figures 2 and 4). In Canada, switching from branded valsartan to a non-valsartan ARB increased in Q2-2018 (Supplemental Figure 4), but this trend was not observed in the other countries. In Denmark, switching from the non-recalled generic and branded products to recalled products prior to Q2-2018 was frequent. After Q2-2018, switching rates diminished.

## DISCUSSION

Our findings suggest that many patients received valsartan products with nitrosamine impurities, at the time when there was no knowledge of the nitrosamine impurities. In all three countries, recalled-tested and recalled valsartan products had the largest share of the valsartan market. Despite widespread use of the products with nitrosamine impurities, duration of use was typically very short. The mean duration of use was around 4 months in the US and Denmark. In Canada, the mean duration of use was slightly longer, at about 8 months. Further, about 75% of the affected valsartan episodes were not longer than 8 months in US and Denmark and 12 months in Canada. This translates to a maximum duration of exposure of 1.5% (12 months out of 840 months (70 years)) of the lifetime exposure for most cancer risk assessments. Our study also revealed that despite availability of non-recalled valsartan products, the preference was to switch to non-valsartan ARB after the recall notices were issued in July 2018.

Nitrosamines, NDMA and NDEA are classified as probable carcinogens for humans given the limited evidence of carcinogenicity in humans but established evidence in animal studies.^5^ NDMA and NDEA are suspected to have both local and systemic carcinogenic potential following activation to diazonium ions (methyldiazonium, ethlydiazonium).^13^ These diazonium ions are precursors of reactive electrophilic carbenium ions, which directly react with DNA to form stable adducts (a segment of the DNA bound to a carcinogenic chemical). Adducts that are not removed by the cell can cause mutations that may give rise to cancer.^14^ As such it is expected that chronic exposure to nitrosamines is necessary for carcinogenesis. With regards to types of cancers, the EMA’s ad-hoc expert group on nitrosamine impurities in human medicinal products^15^ concluded that the liver may be the most impacted organ due to activation of nitrosamines during first pass hepatic metabolism. Activated nitrosamines are very reactive and are unlikely to be released into the circulation. However, many other sites, including the gastrointestinal tract, are metabolically competent and can also activate nitrosamines.^16,17^ Based on the duration of exposure found in our study, a marked increased risk of liver or the gastrointestinal cancer from the nitrosamine impurity is unlikely. An observational study conducted to examine risk of these cancers also found no increased risk for cancers with valsartan products.^18^

Our study has several strengths. It is the first observational study to date to characterize the extent of exposure to valsartan products with nitrosamine impurities in three countries. It covers the entire period when the valsartan products with nitrosamine impurities were in circulation, allowing for the complete capture of nitrosamine exposure from valsartan products described within the study databases. The use of dispensing claims data means that patients filled their prescriptions, though the actual ingestion of these products cannot be confirmed.

## LIMITATIONS

Within the recalled-tested products, it was likely that all products lots for the assigned NDC were affected. However, for the recalled products, only selected lots are indicated on the recalled list were affected by the contamination. For recalled products, our exposure algorithm was unable to identify specific affected lots. Thus, the proportion of recalled valsartan episodes may be overestimated. We were also unable to identify a date when all valsartan products with nitrosamine impurities were no longer on the market. To estimate exposure, we assumed that valsartan products with nitrosamine impurities were no longer on market by December 2018, but we acknowledge that this may not have been the case across the board. Another limitation relates to potential additional contamination exposure from switching to non-valsartan ARBs that were affected prior to issuance of the recall notices. It is possible that prior to the recall dates valsartan users switched to other affected ARBs and could have had additional exposure to nitrosamine impurities. However, we were unable to evaluate this possibility as the date of first introduction of the nitrosamine impurities for the other ARBs was not well characterized at the time of the analysis. Lastly, we acknowledge that there could be other sources of nitrosamine exposure from water and food, which may also affect cancer risk, and these were not considered in this study.

## CONCLUSION

Between 2012 and 2018, recalled generic valsartan products had the largest share of the valsartan market in the US, Canada, and Denmark. Despite widespread use, the duration of use of recalled products was relatively short and likely does not pose an elevated risk of nitrosamine-induced cancer. However, since products with nitrosamine impurity could have been on the market over a six-year period, patients potentially exposed to these products for longer duration could have a different risk of cancer.

## Data Availability

Data cannot be shared with external stakeholders due to confidentiality agreements.

## Acknowledgments, Sources of Funding, & Disclosures

## a) Acknowledgements

Many thanks are due to those who participated in this project:

The Canadian Network for Observational Drug Effect Studies (CNODES), a collaborating center of the Drug Safety and Effectiveness Network (DSEN). This study was made possible through data-sharing agreements between the CNODES member research centers and the respective provincial governments of Manitoba, Nova Scotia, Ontario, and Saskatchewan.

U.S. Data Partners who provided data used in the analysis: CVS Health (Aetna), Bell, PA; Carelon Research/Elevance Health, Wilmington, DE; Duke University School of Medicine, Department of Population Health Sciences, Durham, NC, through the Centers for Medicare and Medicaid Services which provided data; Humana Healthcare Research Inc., Louisville, KY; Northern California, Division of Research, Oakland, CA; OptumInsight Life Sciences Inc., Boston, MA; Vanderbilt University Medical Center, Department of Health Policy, Nashville, TN, through the TennCare Division of the Tennessee Department of Finance & Administration which provided data.

## Disclaimers

The opinions, results, and conclusions are those of the authors; no endorsement by the provincial governments, data stewards, Health Canada, CADTH or CIHR is intended or should be inferred. The views expressed are the authors’ and not necessarily those of the U.S. Food and Drug Administration, or the U.S. Department of Health and Human Services.

